# *GBA* variants in REM sleep behavior disorder: a multicenter study

**DOI:** 10.1101/19010991

**Authors:** Lynne Krohn, Jennifer A. Ruskey, Uladzislau Rudakou, Etienne Leveille, Farnaz Asayesh, Michele T.M. Hu, Isabelle Arnulf, Yves Dauvilliers, Birgit Högl, Ambra Stefani, Christelle Charley Monaca, Abril Beatriz, Giuseppe Plazzi, Elena Antelmi, Luigi Ferini-Strambi, Anna Heidbreder, Bradley F. Boeve, Alberto J. Espay, Valérie Cochen De Cock, Brit Mollenhauer, Friederike Sixel-Döring, Claudia Trenkwalder, Karel Sonka, David Kemlink, Michela Figorilli, Monica Puligheddu, Femke Dijkstra, Mineke Viaene, Wolfgang Oertel, Marco Toffoli, Gian Luigi Gigli, Mariarosaria Valente, Jean-François Gagnon, Alex Desautels, Jacques Y. Montplaisir, Ronald B. Postuma, Guy A. Rouleau, Ziv Gan-Or

## Abstract

**Objective:** To study the role of *GBA* variants in the risk for isolated rapid-eye-movement (REM)-sleep behavior disorder (iRBD) and conversion to overt neurodegeneration.

**Methods:** A total of 4,147 individuals were included: 1,061 iRBD patients and 3,086 controls. *GBA* was fully sequenced using molecular inversion probes and Sanger sequencing. We analyzed the effects of *GBA* variants on the risk for iRBD, age at onset (AAO) and conversion rates.

**Results:** *GBA* variants were found in 9.5% of iRBD patients compared to 4.1% in controls (odds ratio [OR]=2.45, 95% CI=1.87–3.22, *p*=1×10^−10^). The estimated OR for mild p.N370S variant carriers was 3.69, 95% CI=1.90–7.14, *p*=3.5×10^−5^, while for severe variant carriers it was 17.55, 95% CI=2.11–145.9, *p*=0.0015. Carriers of severe *GBA* variants had an average AAO of 52.8 years, 7-8 years earlier than those with mild variants or non-carriers (*p*=0.029). Of the *GBA* variant carriers with available data, 52.5% had converted, compared to 35.6% in non-carriers (*p*=0.011), with a trend for faster conversion among severe *GBA* variant carriers. However, the results on AAO and conversion were based on small numbers and should be taken with caution.

**Conclusions:** *GBA* variants robustly and differentially increase the risk of iRBD. The rate of conversion to neurodegeneration is also increased and may be faster among severe *GBA* variant carriers, although confirmation will be required in larger samples. Screening for RBD in healthy carriers of *GBA* variants should be studied as a potential way to identify *GBA* variant carriers who will develop a synucleinopathy in the future.

## Introduction

Isolated rapid-eye-movement (REM)-sleep behavior disorder (iRBD) can be considered a prodromal synucleinopathy since >80% of iRBD patients will eventually convert to an overt neurodegenerative syndrome associated with *α*-synuclein accumulation: Parkinson’s disease (PD), dementia with Lewy bodies (DLB) or multiple system atrophy (MSA),^1^ with a conversion rate of about 6% a year.^2^ For unknown reasons, while some iRBD patients convert rapidly, others can remain free of parkinsonism or dementia for decades.^3, 4^

Variants in the gene encoding for the lysosomal enzyme glucocerebrosidase, *GBA*, are strong and relatively common risk factors for PD ^5, 6^ and DLB,^7^ yet their role in MSA is still not clear.^8-10^ PD patients who carry *GBA* variants, as a group, tend to have higher rates of non-motor symptoms, including RBD, cognitive impairment, hyposmia and autonomic dysfunction.^11^ *GBA* variants can be classified as severe or mild based on the type of Gaucher disease (GD) associated with the variant.^12^ Accordingly, patients with severe *GBA* variants have a higher risk for PD, an earlier average age at onset (AAO)^5^ and faster cognitive decline,^13, 14^ compared to PD patients with mild or no *GBA* variants.

Thus far, only few studies with small sample size have examined the role of *GBA* in iRBD, including studies of 69,^15^ 171^16^ and 265 iRBD patients,^17^ all supporting an association between *GBA* variants and iRBD, but with different risk estimates. In addition, it has been shown that in PD cohorts with available data on probable RBD (pRBD), *GBA* variants are more frequent in the group with pRBD.^17^ However, there are no accurate estimates of the risk for iRBD among *GBA* variant carriers, and there have been no studies separately analyzing severe and mild *GBA* variants. Furthermore, it is not clear whether *GBA* variants affect the rate of conversion from iRBD to overt synucleinopathies, as only two small sample size studies with contradicting results examined this hypothesis. In one, there was no association with the rate of conversion in 8 *GBA* variant carriers with iRBD;^15^ in the other, a faster conversion was shown for 13 *GBA* variant carriers with iRBD compared to non-carriers.^18^

In this study, we analyzed *GBA* variants in a large, multicenter study including 1,061 iRBD patients, more than double the sample than all previous studies combined, and 3,086 controls, all of European origin. We further examined the effects of severe vs. mild *GBA* variants on risk for iRBD, reported AAO of iRBD, and the potential effects on conversion from iRBD to an overt neurodegenerative disease.

## Methods

### Population

The patient population included 1,061 individuals diagnosed with iRBD with video-polysomnography according to the International Classification of Sleep Disorders, version 2 or 3 (ICSD-2/3) criteria.^19^ The recruiting centers and the number of patients from each center are detailed in Table 1. Additional data were available for subsets of samples, including reported AAO of RBD (n=594), age at diagnosis of iRBD (n=599), eventual phenoconversion to an overt neurodegenerative disease (data available for n=584, converted n=218), and rate of phenoconversion (n=217). The data on these variables were collected in 2018. The control population included a total of 3,086 individuals, comprised of 1,317 in-house controls of European origin (confirmed by principal component analysis using available genome-wide association study [GWAS] data compared to data from HapMap v.3 and hg19/GRCh37), and additional 1,769 previously published European controls in which *GBA* was fully sequenced and all the variants were reported (Supplementary Table 1 details these controls and the reported *GBA* variants in each of the papers). The in-house controls had a mean age of 46.5 ± 15.0 years and included 46.6% men, compared to 60.5 ± 9.9 and 81% men in the patients, therefore when analyzing these populations, adjustment for age and sex was performed (see statistical analysis and results).

**Table 1.** *GBA* variants in the participating centers

| Center | N *GBA* variant carriers (%) / N total iRBD patients | *GBA* variants identified | Conversion data available for patients, n | Conversion in *GBA* carriers |
| --- | --- | --- | --- | --- |
| Montreal, Canada | 20 (15%) / 138 | p.N370S – 3<br>p.E326K – 5<br>p.T369M – 7<br>p.L444P – 1<br>p.H255Q – 1<br>p.W378G – 2<br>p.W291X – 1 | 125 | p.E326K – two converted to DLB, one to PD, one to unspecified dementia<br>p.H255Q – one converted to PD<br>p.L444P – one converted to PD<br>p.T369M – three converted to PD, one to MSA, one to unspecified dementia<br>p.W291X – one converted to PD<br>p.W378G – two converted to DLB |
| Innsbruck, Austria | 7 (9%) / 80 | p.E326K – 7 | 69 | p.E326K – one converted to PD |
| Geel, Belgium | 0 (0%) / 9 | - | 8 | - |
| Bologna, Italy | 2 (7%) / 28 | p.E326K – 1<br>p.R2L – 1 | 19 | - |
| Prague, Czech Republic | 3 (6%) / 47 | p.E326K – 1<br>p.T369M – 2 | 46 | p.T369M – one converted to unspecified dementia |
| Nimes, France | 0 (0%) / 5 | - | - | - |
| Paris, France | 14 (6%) / 219 | p.E326K – 8<br>p.N370S – 4<br>p.T369M – 2 | 77 | p.E326K – one converted to PD, one to dementia<br>p.N370S – one converted to PD |
| Montpellier, France<br>(Beau Soleil Clinic) | 5 (19%) / 26 | p.E326K – 2<br>p.N370S – 1<br>p.T369M – 1<br>p.R502C – 1 | 2 | p.R502C – one converted to PD |
| Lille, France | 5 (22%) / 23 | p.E326K – 2<br>p.N370S – 2<br>p.R131L – 1 | 15 | p.N370S – one converted to PD, one to MSA<br>p.R131L – one converted to PD |
| Montpellier, France<br>(CHU Montpellier) | 8 (8%) / 96 | p.E326K – 5<br>p.N370S – 2<br>p.T369M – 1 | 14 | p.T369M – one converted to DLB |
| Marburg, Germany | 0 (0%) / 29 | - | - | - |
| Kessel, Germany | 3 (11%) / 27 | p.E326K – 2<br>p.T369M – 1 | 27 | p.T369M – one converted to DLB |
| Munster, Germany | 0 (0%) / 23 | - | 23 | - |
| Cagliari, Italy | 0 (0%) / 28 | - | 18 | - |
| Udine, Italy | 6 (7%) / 83 | p.E326K – 2<br>p.N370S – 2 | 25 | p.E326K – one converted, type unknown |
|  |  | p.T369M – 1<br>p.D409H – 1 |  | p.N370S – one converted, type<br>unknown<br>p.D409H – one converted, type<br>unknown |
| Milan, Italy | 1 (5%) / 19 | p.T369M – 1 | 19 | none |
| Oxford, UK | 27 (15%) / 181 | p.E326K – 12<br>p.N370S – 6<br>p.N227S – 1<br>p.T369M – 4<br>p.T369M/p.E326K<br>– 1<br>p.D409H – 1<br>p.R2L – 1<br>p.Y212H – 1 | 97 | p.E326K – two converted to PD<br>p.N370S – one converted to PD |

### GBA sequencing and classification of GBA variants

*GBA* was fully sequenced as previously described,^20^ and the full protocol is available upon request. In brief, molecular inversion probes (MIPs) targeting the coding sequence of *GBA* were designed, and next generation sequencing (NGS) was performed post capture. Alignment, variants calling and annotations were done as previously described,^20^ using standard pipeline. Exons 10 and 11 were also sequenced using Sanger sequencing, since the coverage of NGS of these exons was low. Supplementary table 2 details the probes used for the MIPs capture.

Classification of *GBA* variants as severe or mild was performed as previously described,^5, 12^ based on the occurrence of these variants in the severe (type II and type III) and mild (type I) forms of GD. The p.E326K and p.T369M variants, which do not cause GD but have a comparable risk for that of the p.N370S variants in PD,^21, 22^ were therefore included in the mild variant group.

### Statistical analysis

To examine the association between *GBA* variants and risk for iRBD and controls, association tests (chi-square or Fisher exact test), logistic regression adjusted for sex and age, and burden tests were used. To examine the association of *GBA* variants with risk for iRBD comparing all controls, Chi-square or Fisher exact tests were used, since there was no available data on age and sex from the controls collected from the literature to perform adjusted logistic regression. We therefore further performed this association using only our in-house European controls for which data on age and sex were available, using logistic regression model adjusted for age and sex. Of note, having younger controls may result in under-estimation of the risk, as some of the young controls with *GBA* variants may develop iRBD and/or overt neurodegeneration in the future.

Therefore, if the statistical adjustment is not complete, the risk estimations that were calculated could be slightly lower (i.e. false positive results are not likely, rather under-estimated risk is likely). We also performed burden tests using the R package SKAT. Association with AAO and specific types of *GBA* variants (severe or mild) was tested using the non-parametric Kruskal-Wallis test since the group of severe *GBA* variants included only five patients. The association with conversion was tested using a chi-square test for the total number of conversions, and Kaplan-Meier survival analysis was performed to examine the rate of conversion. All statistical analyses were performed using R or SPSS v24 (IBM).

### Standard Protocols Approvals, Registrations, and Patient Consents

All study participants signed informed consent forms, and the study protocol was approved by the institutional review boards.

### Data availability statement

Anonymized data will be shared by request from any qualified investigator.

## Results

### *GBA* variants are associated with increased risk of iRBD, with differential effects of severe and mild variants

The variants in *GBA* identified in each of the participating centers are detailed in Table 1, with a total of 17 distinct variants found in patients and controls (Table 2). Supplementary Table 1 details the variants found in each of the previously published control populations. Out of 1,061 iRBD patients, 101 *GBA* variant carriers (9.5%) were identified, compared to 126 out of 3,086 (4.1%) controls (Table 2, OR=2.45, 95% CI 1.87–3.22, *p*=1×10^−10^). We repeated this analysis using a logistic regression model adjusted for age and sex using the controls with available data (n=1,317), which yielded very similar results (OR=2.12, 95% CI 1.34-3.36, *p*=0.001). Burden tests using the R package SKAT also yielded similar results (*p*=2.6×10^−6^ using the in-house controls and *p*=1.7×10^−12^ using all controls). Similar to previous observations in PD, different *GBA* variants have different effects on the risk for iRBD. The mild p.N370S variant was found in 20 iRBD patients (1.9%) compared to 16 (0.5%) in the controls (OR=3.69, 95% CI 1.90–7.14, *p*=3.5×10^−5^), while severe variants (p.L444P, p.D409H, p.W291X, p.H255Q and p.R131L) were found in six (0.6%) iRBD patients and in one (p.L444P, 0.03%) control (OR 17.55, 95% CI 2.11–145.9, *p*=0.0015). Of the two polymorphisms known to be risk factors for PD, p.E326K and p.T369M, only p.E326K was associated with iRBD (4.4% vs. 1.5% in patients and controls, OR 3.2, 95% CI 2.12-4.84, *p*=6×10^−9^), and the carrier frequency of p.T369M was only slightly elevated in iRBD but not statistically significant (1.9% vs. 1.7%, OR 1.13, 95% CI 0.68-1.89, *p*=0.6). The carrier frequencies of the p.N370S, p.E326K and p.T369M variants in gnomAD (https://gnomad.broadinstitute.org/) European population are 0.4%, 2.4% and 1.9% respectively, similar to the frequencies in our controls.

**Table 2.** *GBA* variants in iRBD patients and controls

| *GBA* variant<sup>a</sup> | iRBD patients,<br>n (%) | All controls, n<br>(%) | In-house<br>controls, n (%) | Literature<br>controls, n (%) |
| --- | --- | --- | --- | --- |
|  | n=1,061 | n=3,086 | n=1,317 | n=1,769 |
| Heterozygous |  |  |  |  |
| p.R2L | 2 (0.2%) | 2 (0.06%) | 1 (0.08%) | 1 (0.06%) |
| p.K79M | - | 1 (0.03%) | - | 1 (0.06%) |
| p.R131L | 1 (0.1%) | - | - | - |
| p.Y212H | 1 (0.1%) | - | - | - |
| p.N227S | 1 (0.1%) | - | - | - |
| p.H255Q | 1 (0.1%) | - | - | - |
| p.W291X | 1 (0.1%) | - | - | - |
| p.E326K | 47 (4.4%) | 45 (1.4%) | 18 (1.4%) | 27 (1.5%) |
| p.T369M | 20 (1.9%) | 54 (1.7%) | 35 (2.7%) | 19 (1.1%) |
| p.N370S | 20 (1.9%) | 16 (0.5%) | 10 (0.8%) | 6 (0.3%) |
| p.W378G | 2 (0.2%) | 1 (0.03%) | 1 (0.08%) | - |
| p.E388K | - | 3 (0.09%) | - | 3 (0.17%) |
| p.D409H | 2 (0.2%) | - | - | - |
| p.L444P | 1 (0.1%) | 1 (0.03%) | - | 1 (0.06%) |
| p.V460L | - | 2 (0.06%) | - | 2 (0.11%) |
| p.T482K | - | 1 (0.03%) | - | 1 (0.06%) |
| p.R502C | 1 (0.1%) | - | - | - |
| Homozygous / compound<br>heterozygous |  |  |  |  |
| p.E326K/p.T369M | 1 (0.1%) | - | - | - |
| Total | 101 (9.5%) | 126 (4.1%) | 65 (4.9%) | 61 (3.4%) |

<sup>a</sup> Variants nomenclature is according to the nomenclature typically used for *GBA* variants, of the active enzyme (497 amino acids) after the removal of the 39 amino acids of the leader peptide.
iRBD, isolated REM-sleep behavior disorder; n, number

### Estimated age at onset of iRBD may be affected by the type of *GBA* variant

AAO as reported by the patients could be an unreliable estimate, and data was not available for all patients. Therefore, the following results should be considered with caution. Carriers of the severe *GBA* variants had an average AAO of 52.8 years ± 2.8 years (data was available for five out of six patients with a severe *GBA* variant), carriers of all other variants had an average AAO of 59.7 ± 9.6 years (data was available for 58 patients), and non-carriers of *GBA* variants had an average AAO of 60.6 ± 9.9 years (data was available for 531 patients). Since in the severe variants group there were only five patients, the non-parametric Kruskal-Wallis test was performed, demonstrating a possible association with the type of variant (χ^2^=7.083, df=3, *p*=0.029).

### Do *GBA* variants affect the rate of conversion of iRBD to overt neurodegenerative diseases?

Data on conversion of iRBD was available for 59 *GBA* variant carriers and 525 non-carriers of *GBA* variants. Of the *GBA* variant carriers, 31 (52.5%) had converted, and in non-carriers 187 (35.6%) had converted (*p*=0.011). Data on time from iRBD diagnosis to phenoconversion or last follow-up was available for 29 *GBA* variant carriers and for 276 non-carriers. Kaplan-Meier survival analysis suggested that *GBA* variant carriers progressed faster but the difference with non-carriers of *GBA* variants was not statistically significant (Figure 1A). When severe *GBA* variant carriers were compared to mild *GBA* carriers and non-carriers, a possible association was demonstrated (Figure 1B; Breslow *p*=0.017, Tarone-Ware *p*=0.051, log-rank *p*=0.24). However, these results should be interpreted with caution for several reasons: a) They include the cohort from Montreal in which it was previously reported that *GBA* variants are associated with rate of conversion, but it does not include the negative study from Barcelona (data could not be shared). b) The results are based on a small number of variant carriers (four patients with a severe *GBA* variant, 25 with other *GBA* variants). Therefore, larger studies will be required to conclusively determine whether *GBA* variants are associated with the rate of phenoconversion.

**Figure 1.**
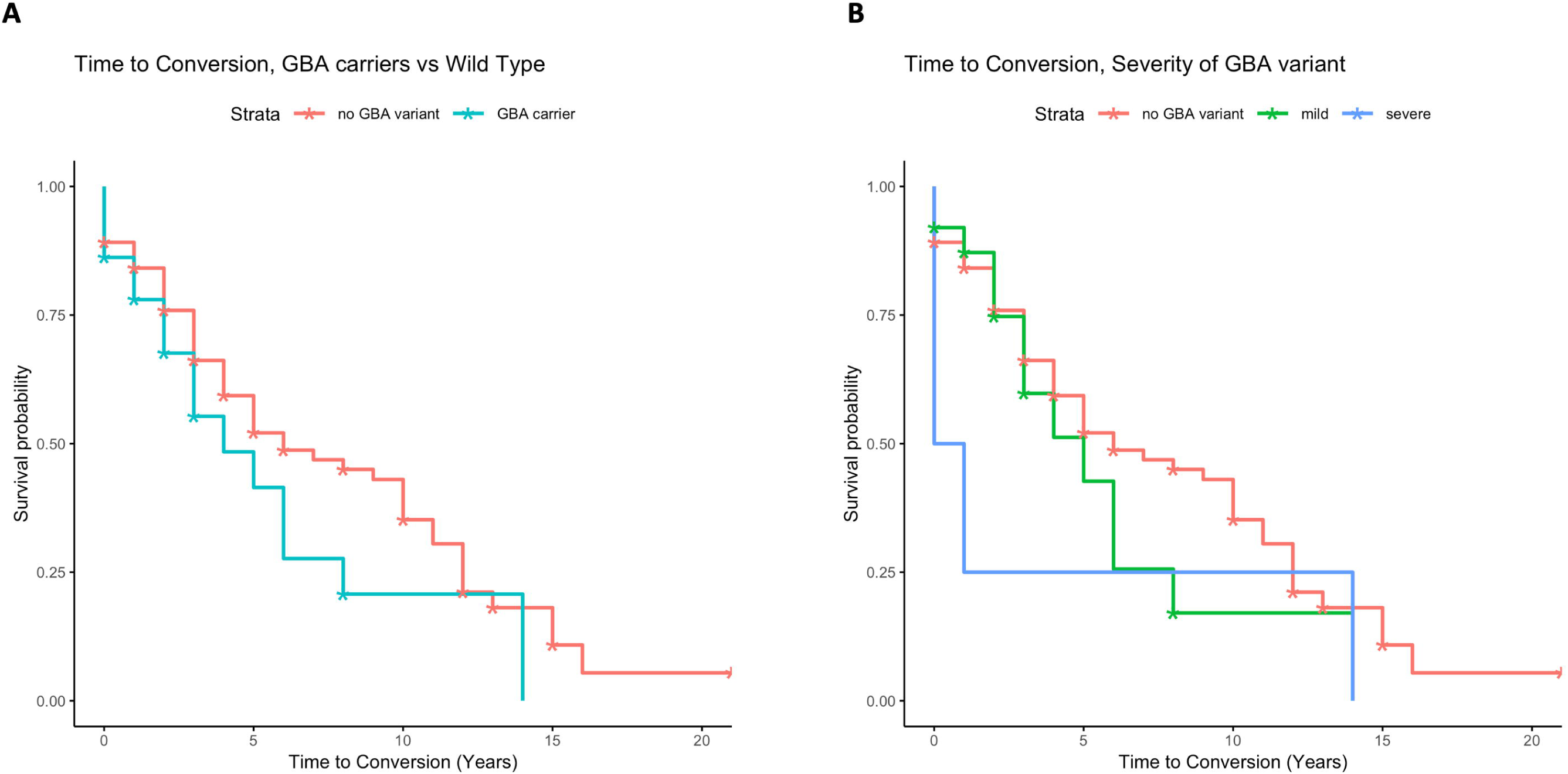
Conversion to overt neurodegenerative disease in iRBD patients with and without *GBA* variantss. A) Survival plot comparing *GBA* variant carriers (green) and non-carriers (blue) from diagnosis until conversion or recent follow-up. Log-Rank *p*=0.13, Breslow *p*=0.32, Tarone-Ware *p*=0.22. B) Survival plot comparing carriers of severe *GBA* variants, mild/other *GBA* variants (green) and non-carriers (blue). Log-Rank *p*=0.24, Breslow *p*=0.017, Tarone-Ware *p*=0.051.

## Discussion

Our results confirm the association between *GBA* variants and increased risk for iRBD, and suggest that severe and mild *GBA* variants have differential effects on risk, similar to previous reports in PD.^5^ Furthermore, our results suggest that iRBD patients with severe *GBA* variants may have earlier AAO, and may convert faster to overt neurodegenerative disease. However, the results on AAO and conversion should be considered as preliminary only and with caution, due to several limitations discussed below.

Three previous small sample size studies have examined the association between *GBA* variants and iRBD.^15-17^ Two of these studies included full sequencing of the gene,^15, 17^ and the third only examined two specific variants (p.N370S and p.L444P).^16^ Due to their size, analyses of specific variants or types of variants such as severe or mild were not possible. The current study includes two of the previously published cohorts,^16, 17^ and additional cohorts of European ancestry. With the larger sample size accrued, we were able to demonstrate a much larger risk in carriers of severe *GBA* variants. However, given the small numbers of these variants and the wide range of the confidence interval, the risk estimates may be different in future, larger studies. Nevertheless, the current results are in line with previous results from PD, which clearly demonstrated similar relationships between severe and mild *GBA* variants and risk for PD.^5^ Furthermore, previous studies have also suggested that the type of *GBA* variants may affect PD progression,^13, 14^ which is further supported by our preliminary findings on AAO and conversion of iRBD.

In recent years, it has been demonstrated that the two coding variants, p.E326K and p.T369M, which do not cause GD, are risk factors for PD.^21-23^ In DLB, the association between p.E326K and risk for the disease is clear, yet it is still unclear whether p.T369M is a risk factor for DLB. Only a few studies that examined p.T369M in DLB have been performed, and in most of them there was no association between p.T369M. A multicenter study which included over 700 DLB patients reported lack of association, and in a GWAS with over 1,700 DLB patients, only the p.E326K variant was reported to be associated with the disease.^7^ Conversely, recent data from 556 DLB patients did suggest an association.^24^ The lack of association in the current study in iRBD may also provide further support for lack of association of p.T369M with iRBD and DLB. However, it is important to keep in mind that the association of this variant with PD was only reported in much larger studies,^21, 22^ due to its lower effect on risk compared to other *GBA* variants. Only much larger studies can determine conclusively whether p.T369M is associated with iRBD and DLB. Furthermore, there was a large difference between the frequency of p.T369M in our in-house controls (2.7%) and the controls from the literature (1.1%), perhaps due to population structure. However, the combined frequency (1.7%) is comparable to that seen in the gnomAD European population (1.9%), rendering our results for this variant as likely unbiased.

Our study has several limitations. The possible association between *GBA* variants and rate of conversion reported here, although potentially very interesting, should be taken with caution. As detailed in the results, they are based on a small number of patients with severe *GBA* variants, and include samples that were previously included in a study suggesting a positive association between *GBA* variants and rate of conversion. An additional potential limitation is that the measured duration from age at diagnosis or iRBD to conversion might not reflect the actual length of disease duration, as patients can remain unaware for many years about their dream enactment behaviors, especially if they do not have a bed partner or if they do not have very active or violent dreams. The small number of severe *GBA* variants is also a limitation in the risk analysis, as it created a wide confidence interval. However, since the effect of severe vs. mild variants is in line with previous studies in PD, it is likely that these risk estimates for iRBD are overall correct, yet the precise estimate might change in future, larger studies.

The mechanisms underlying the association between *GBA* variants, the enzyme encoded by *GBA*, glucocerebrosidase (GCase), and the development of neurodegeneration are still unknown.^11^ Several mechanisms have been proposed, including interaction of GCase substrates with a-synuclein which may lead to its accumulation,^25^ changes in the lysosomal membrane composition which may lead to reduced autophagy and mitophagy,^26, 27^ accumulation of misfolded GCase and endoplasmic reticulum stress,^28^ and others. The association with iRBD may suggest that studying these mechanisms in non-dopaminergic neuronal models which are involved in RBD could lead to new discoveries and better understanding of these potential mechanisms.

To conclude, our results clearly demonstrate that *GBA* variants are associated with increased risk for iRBD, and suggest that severe and mild *GBA* variants may have differential effects on the risk, and possibly on AAO of iRBD and conversion to overt neurodegenerative disease. Due to the limitations mentioned above, the latter associations should be considered as preliminary, with additional, larger studies on *GBA* in iRBD required to confirm or refute them. One important implication of the association between *GBA* variants and iRBD, is the possibility to perform screening for iRBD in healthy *GBA* variant carriers. This may allow for even earlier detection of prodromal neurodegeneration, and could be especially useful when home detection of iRBD will be made possible.

## Data Availability

Anonymized data is available upon request from the corresponding author

## Acknowledgements

We thank the patients and control subjects for their participation in this study. This work was financially supported by Parkinson Canada, the Michael J. Fox Foundation, the Canadian Consortium on Neurodegeneration in Aging (CCNA), the Canadian Glycomics Network (GlycoNet), the Canada First Research Excellence Fund (CFREF), awarded to McGill University for the Healthy Brains for Healthy Lives (HBHL) program, the Canadian Institutes for Health Research (CIHR), Weston Foundation, Fonds de recherche du Québec - Santé (FRQS) Chercheurs-boursiers. The Oxford Discovery study is funded by the Monument Trust Discovery Award from Parkinson’s UK and supported by the National Institute for Health Research (NIHR) Oxford Biomedical Research Centre based at Oxford University Hospitals NHS Trust and University of Oxford, the NIHR Clinical Research Network and the Dementias and Neurodegenerative Diseases Research Network (DeNDRoN). We thank Helene Catoire, Sandra B. Laurent and Dan Spiegelman for their assistance. JFG holds a Canada Research Chair in Cognitive Decline in Pathological Aging. WO is Hertie Senior Research Professor, supported by the Charitable Hertie Foundation, Frankfurt/Main, Germany. EAF holds a Canada Research Chair (Tier 1) in Parkinson Disease. GAR holds a Canada Research Chair in Genetics of the Nervous System and the Wilder Penfield Chair in Neurosciences. ZGO is supported by the Fonds de recherche du Québec - Santé (FRQS) Chercheurs-boursiers award given in collaboration with Parkinson Quebec, and is a Parkinson Canada New Investigator awardee.

## APPENDIX 1

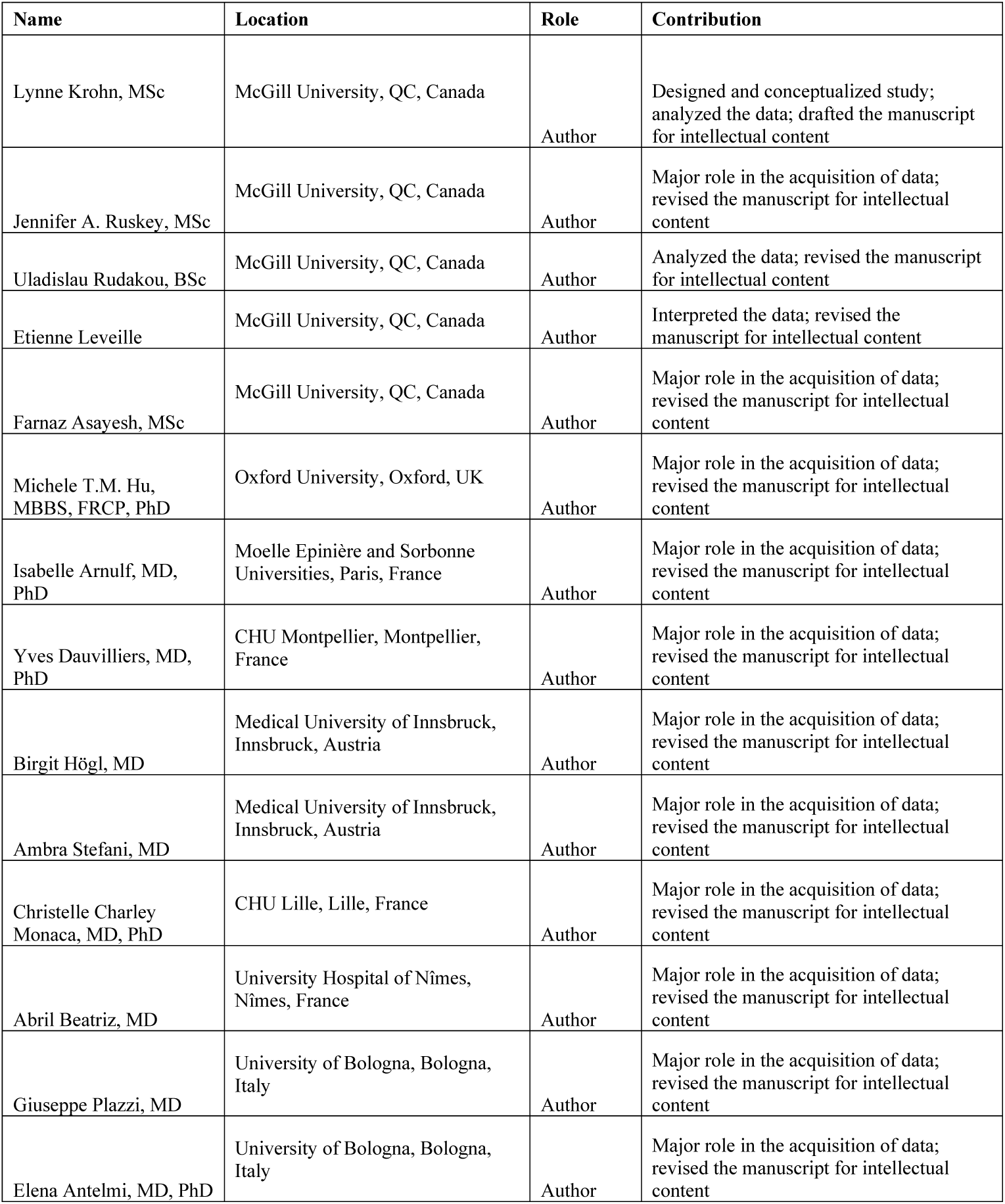
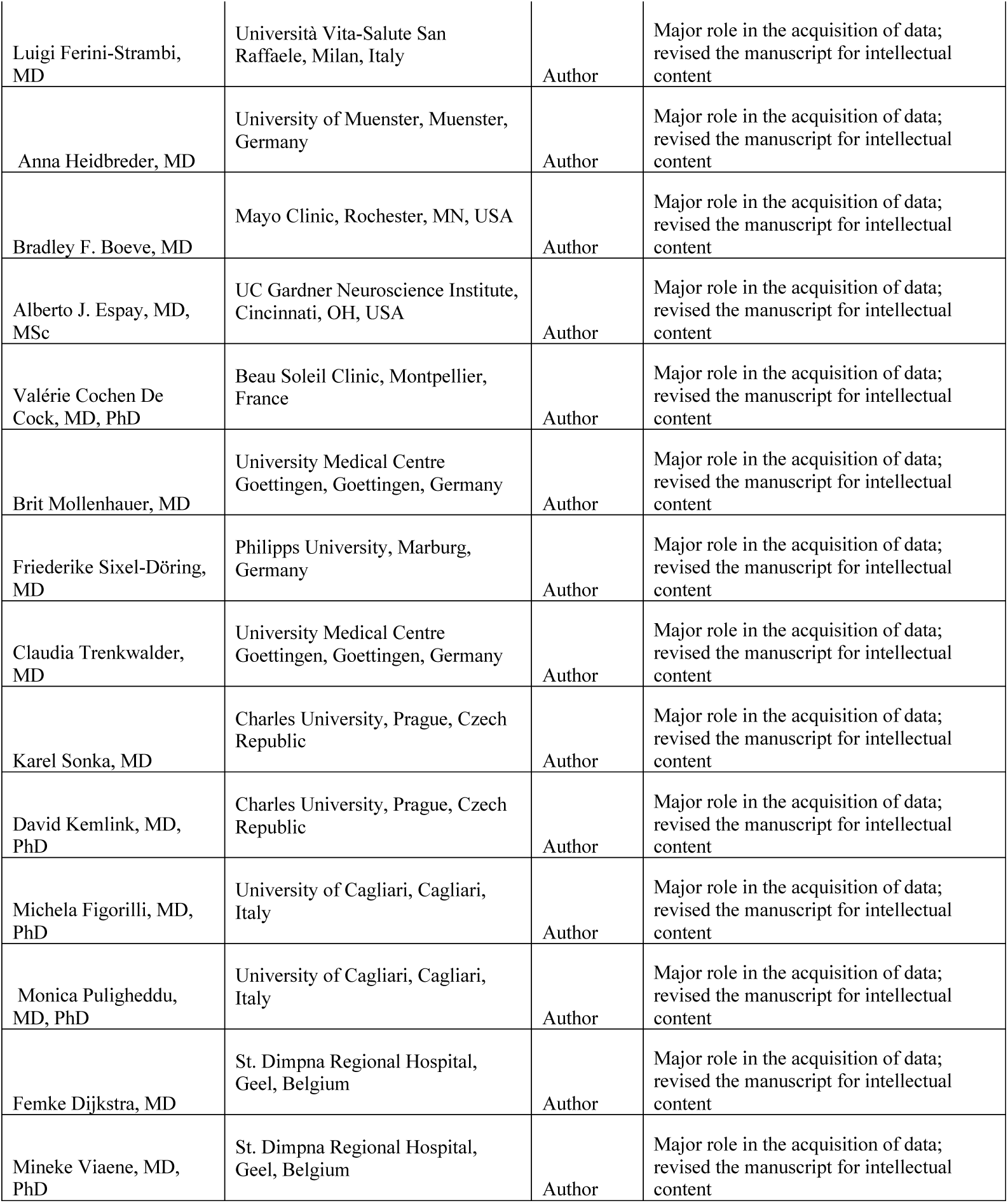
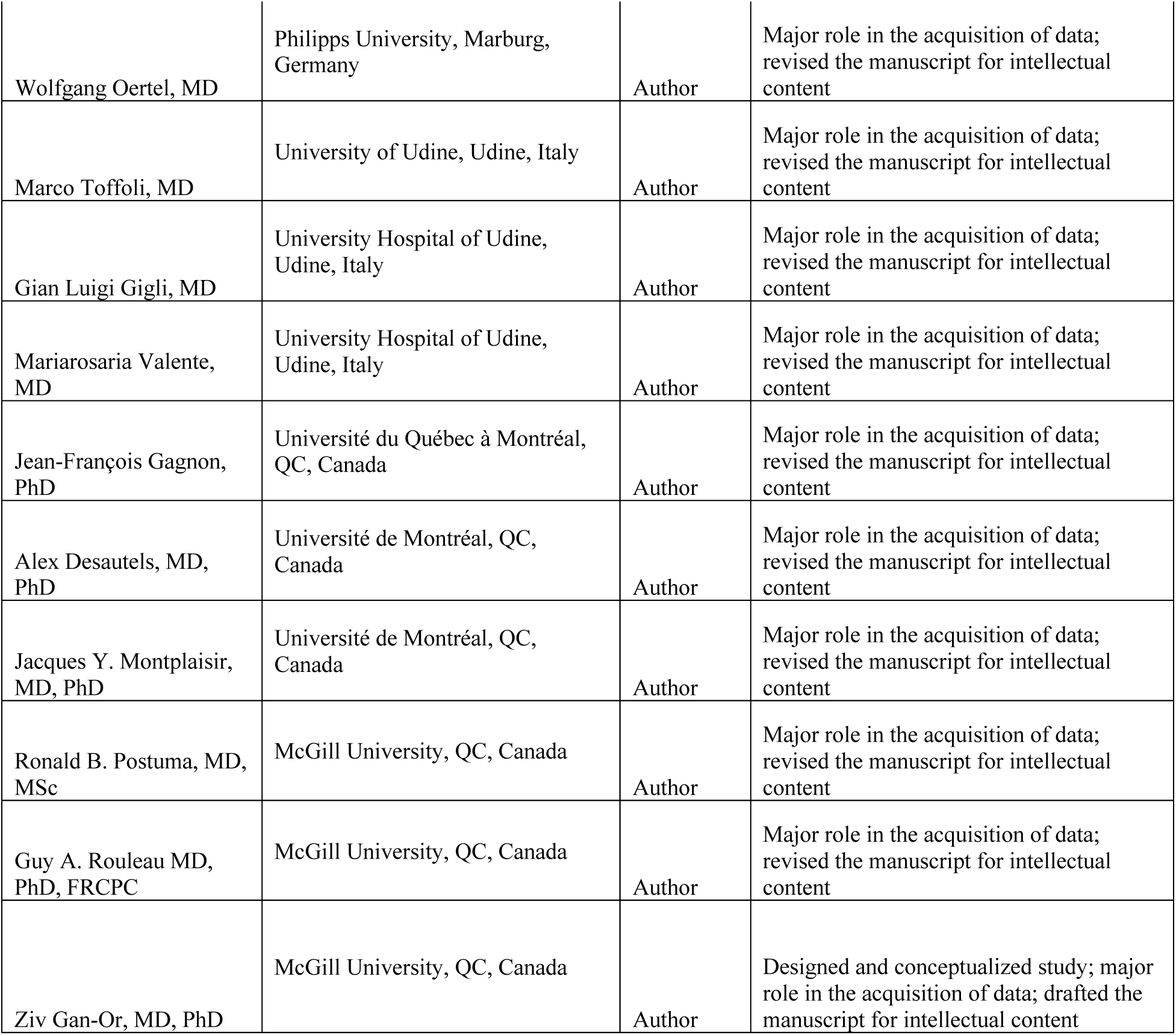

## Notes

### Competing Interest Statement

Lynne Krohn – Reports no conflict of interests
Jennifer A. Ruskey – Reports no conflict of interests
Uladislau Rudakou - Reports no conflict of interests
Etienne Leveille – Reports no conflict of interests
Farnaz Asayesh – Reports no conflict of interests
Michele T.M. Hu – Received consultancy fees from Roche and Biogen Pharmaceuticals
Isabelle Arnulf – Received fees for speaking engagements from UCB pharma, consultancy for Roche, Novartis, and Ono Pharma.
Yves Dauvilliers – Reports no conflict of interests
Birgit Högl – Received consultancy fees from Axovant, benevolent Bio, Takeda, Roche, ono, and received speaker honoraria from Eli Lilly, Mundipharma, UCB, Abbvie, Inspire, Lundbeck.
Ambra Stefani – Reports support for research from Axovant
Christelle Charley Monaca – Reports no conflict of interests
Abril Beatriz – Reports no conflict of interests
Giuseppe Plazzi – Reports no conflict of interests
Elena Antelmi – Reports no conflict of interests
Luigi Ferini-Strambi – Reports no conflict of interests
Anna Heidbreder – Received honoraria for lectures from UCB, Bioprojet, Servier, Medice and consultancy fees from UCB.
Bradley F. Boeve – Dr. Boeve has served as an investigator for clinical trials sponsored by Biogen and Alector. He serves on the Scientific Advisory Board of the Tau Consortium. He receives research support from the NIH, the Mayo Clinic Dorothy and Harry T. Mangurian Jr. Lewy Body Dementia Program, the Little Family Foundation, and the LBD Functional Genomics Program.
Alberto J. Espay – Received grant support from the NIH and the Michael J Fox Foundation; personal compensation as a consultant/scientific advisory board member for Abbvie, Adamas, Acadia, Acorda, Neuroderm, Neurocrine, Impax/Amneal, Sunovion, Lundbeck, Osmotica Pharmaceutical, and USWorldMeds; publishing royalties from Lippincott Williams & Wilkins, Cambridge University Press, and Springer; and honoraria from USWorldMeds, Lundbeck, Acadia, Sunovion, the American Academy of Neurology, and the Movement Disorders Society.
Valérie Cochen De Cock – Reports no conflict of interests
Brit Mollenhauer – Has received honoraria for consultancy from Roche, Biogen, UCB and Sun Pharma Advanced research Company. BM is member of the executive steering committee of the Parkinson Progression Marker Initiative and PI of the Systemic Synuclein Sampling Study of the Michael J. Fox Foundation for Parkinson’s Research and has received research funding from the Deutsche Forschungsgemeinschaft (DFG), EU (Horizon2020), Parkinson Fonds Deutschland, Deutsche Parkinson Vereinigung and the Michael J. Fox Foundation for Parkinson’s Research.
Friederike Sixel-Döring – Honoraria for lectures from Abbott, Desitin, Grünenthal, Licher MT, STADA Pharm, UCB. Seminar fees from Boston Scientific, Licher MT. Serves on an advisory board for STADA Pharm. No conflict of interest with the presented study.
Claudia Trenkwalder – Honoraria for lectures from UCB, Grünenthal, Otsuka and consultancy fees from Britannia Pharmaceuticals and Roche. 
Karel Sonka – Reports no conflict of interests
David Kemlink – Reports no conflict of interests
Michela Figorilli – Reports no conflict of interests
Monica Puligheddu – Reports no conflict of interests
Femke Dijkstra – Reports no conflict of interests
Mineke Viaene – Reports no conflict of interests
Wolfgang Oertel – Reports no conflict of interests related to the study. He received consultancy or speaker fees from Adamas, Abbvie, Desitin, Novartis and Roche.
Marco Toffoli – Reports no conflict of interests
Gian Luigi Gigli – Reports no conflict of interests
Mariarosaria Valente – Reports no conflict of interests
Jean-François Gagnon – Reports research funding from the Canadian Institutes for Health Research (CIHR).
Alex Desautels – Received grants from Flamel Ireland, Pfized, Biron, Canopy Growth, as well as fees for speaking engagements from Biogen and from consultancy from UCB pharma.
Jacques Y. Montplaisir – Reports no conflict of interests
Ronald B. Postuma – Reports grants and Fonds de la Recherche en Sante, as well as grants from the the Canadian Institute of Health Research, The Parkinson Society of Canada, the Weston-Garfield Foundation, the Michael J. Fox Foundation, and the Webster Foundation, as well as personal fees from Takeda, Roche, Teva Neurosciences, Novartis Canada, Biogen, Boehringer Ingelheim, Theranexus, GE HealthCare, azz Pharmaceuticals, Abbvie, Jannsen, Otsuko, Phytopharmics, and Inception Sciences
Guy A. Rouleau – Reports no conflict of interests
Ziv Gan-Or – Received consultancy fees from Lysosomal Therapeutics Inc. (LTI), Idorsia, Prevail Therapeutics, Inceptions Sciences (now Ventus), Denali and Deerfield. 

